# Estimating the distribution of COVID-19-susceptible, -recovered, and -vaccinated individuals in Germany up to April 2022

**DOI:** 10.1101/2022.04.19.22274030

**Authors:** Benjamin F. Maier, Annika H. Rose, Angelique Burdinski, Pascal Klamser, Hannelore Neuhauser, Ole Wichmann, Lars Schaade, Lothar H. Wieler, Dirk Brockmann

## Abstract

After having affected the population for two years, the COVID-19 pandemic has reached a phase where a considerable number of people in Germany have been either infected with a SARS-CoV-2 variant, vaccinated, or both. Yet the full extent to which the population has been in contact with either virus or vaccine remains elusive, particularly on a regional level, because (a) infection counts suffer from under-reporting, and (b) the overlap between the vaccinated and recovered subpopulations is unknown. Since previous infection, vaccination, or especially a combination of both reduce the risk of severe disease, a high share of individuals with SARS-CoV-2 immunity lowers the probability of severe outbreaks that could potentially overburden the public health system once again, given that emerging variants do not escape this reduction in susceptibility. Here, we estimate the share of immunologically naïve individuals by age group for each of the 16 German federal states by integrating an infectious disease model based on weekly incidences of SARS-CoV-2 infections in the national surveillance system and vaccine uptake, as well as assumptions regarding under-ascertainment. We estimate a median share of 7.0% of individuals in the German population have neither been in contact with vaccine nor any variant as of March 31, 2022 (quartile range [3.6%– 9.8%]). For the adult population at higher risk of severe disease, this figure is reduced to 3.5% [1.3%–5.5%] for ages 18–59 and 4.3% [2.7%–5.8%] for ages 60 and above. However, estimates vary between German states mostly due to heterogeneous vaccine uptake. Excluding Omicron infections from the analysis, 16.1% [14.0%–17.8%] of the population in Germany, across all ages, are estimated to be immunologically naïve, highlighting the large impact the Omicron wave had until the beginning of spring in 2022.

## I INTRODUCTION

The COVID-19 pandemic caused by the rapid global dissemination of the SARS-CoV-2 virus and its respective variants has led to a large number of infections worldwide [1]. In Germany, around 21.4 million infections have been reported as of the end of March 2022. Moreover, a large part of the population has received a primary vaccination series with one of the available COVID-19 vaccines (mRNA-vaccine by BioNTech or Moderna, or a vector-based vaccine by AstraZeneca or Janssen) [2]. The national COVID-19 vaccination campaign began at the end of 2020 by targeting older adults, residents of nursing homes, and healthcare workers, then shifting focus to younger adults [3]. In August 2021, a recommendation to vaccinate adolescents aged 12-17 was issued [4] and since December 2021, children aged 5-11 years are recommended to receive a vaccination if underlying medical conditions put them at increased risk for severe disease [5]. In Germany, recovered individuals are advised not to receive a COVID-19 vaccination until 6 months [6], or 3 months [7] have passed after infection. At the time of analysis, booster vaccinations have been recommended for all persons aged 11 years and older [8, 9]. A central factor that will determine how the pandemic progresses in Germany in the near future is the number of people still immunologically naïve to infection, i.e. that have neither been in contact with the virus or any of its variants nor a vaccine against them. In Germany, several serological studies have been conducted [10, 11], but none that extend into the time of the Omicron waves, particularly with respect to children. Therefore, we choose a mathematical modeling approach here to estimate the number of immunologically naïve individuals in order to facilitate informed decisions with regard to the upcoming pandemic situation in the fall of 2022.

To estimate the number of people that have been in contact with either virus or vaccine, one might simply summate the number of vaccinations and the number of reported infections. However, doing so ignores the fact that (a) a considerable number of vaccinated people have suffered from additional breakthrough infections (taking into account both asymptomatic and symptomatic infections herein) [12], (b) a substantial number of previously infected people have chosen to be vaccinated in accordance with national recommendations [13–15], (c) some individuals have suffered from multiple infections [16], and (d) the exact extent of the total number of infections as compared to the reported number of infections is unknown because (i) asymptomatic infections are less likely to be identified and reported in the national surveillance system and (ii) underascertainment varies regionally [17, 18]. In order to estimate the overlap between the vaccinated and recovered subpopulations, one may assume that the probability of any recovered individual to be vaccinated is proportional to the probability of any individual to be vaccinated. However, this largely ignores (i) the heterogeneous dynamics of the spreading disease and vaccination campaigns, and (ii) that vaccinated individuals are less likely to suffer from an infection than unvaccinated individuals [19]. Here, we introduce modeling approaches that are devised to meet the aforementioned conditions and use them to estimate the distribution of immunologically naïve, (in the infectious disease modeling context called “fully susceptible” hereafter), recovered, and vaccinated individuals in Germany, taking into account regional and age differences. We find that although the percentage of the adult population in Germany that remains fully susceptible is expected to be in the single digits, the share of unaffected children may be considerably larger. Due to heterogeneities in vaccine uptake across German states, these values may differ by region. Our analysis cannot answer questions regarding the quality of achieved immunity against infection or disease, because we consider neither waning of immunity nor the emergence of variants with immune evasive properties, which is difficult to predict [20].

## II. METHODS

We partition the population into *n*_*G*_ = 16 regions corresponding to the German states and *n*_*A*_ = 5 age groups corresponding to ages “00-04” (infants), “05-11” (children), “12-17” (adolescents), “18-59” (adults), “60+” (elderly), chosen in accordance with the population structure of publicly available vaccination data [2], i.e. into 80 subpopulations. To obtain nation-wide counts of individuals in age groups, we sum the respective results over all regions, to obtain counts of individuals for all ages, we sum over all age groups. To obtain an age-independent, nation-wide result, we sum over all ages and all regions.

As we are, first and foremost, interested in estimating the proportion of individuals *S*_∞_ ≡ *S*(*t* = *t*_max_) that can be considered to be fully susceptible towards infection with any SARS-CoV-2 variant per region and age group, we report a simplified model here that captures the main ideas and gives the same results for *S*(*t*) as the full model which is reported in the Appendix (see App. A 1).

We consider the population of size *N* (an age group in a region) to be composed of susceptible (*S*), infected/recovered (*I*), infected/recovered but eligible for reinfection or vaccination (*Y*), vaccinated (*V*), and boostered (*B*) individuals, assuming that the population count is constant over two years such that *N* = *S* + *I* +*Y* +*V* + *B* = const.

The central problem of estimating *S*_∞_ is to determine the overlap between recovered and vaccinated subpopulations. Given that the cumulative number of unvaccinated infected *R*_∞_ and the number of cumulative vaccinated individuals *V*_∞_ is known, one may naively assume that the probability that an infected person that was initially unvaccinated is vaccinated later on is proportional to the probability that any person in the population is vaccinated, which is given as *p* = *V*_∞_/*N*. Then, the cohort size of unvaccinated and not yet infected individuals is *S*_∞_ = *N* − (1 −*V*_∞_/*N*) *R*_∞_ −*V*_∞_. However, this largely ignores the time course of infections and vaccinations, with incidence and daily vaccinations peaking at different time points, with a large number of infections occurring after the peak in vaccinations. Hence, one may assume instead that when a person becomes vaccinated at time *t*, the probability that this person was already infected is proportional to the number of infected/recovered individuals at time *t* that are eligible for vaccination as *p* = *Y* /(*S* +*Y*). With incidence rates of *a*_*ϕ*_*ϕ*(*t*) (new unvaccinated cases per day) and vaccination rates of *a*_*β*_*β*_*S*_ (*t*) (new vaccinations per day) obtained from data, we assume that the count of individuals in the respective states evolves dynamically as

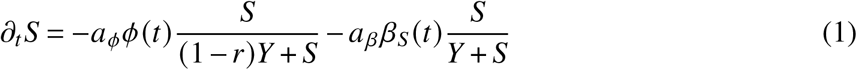

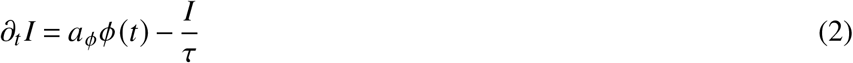

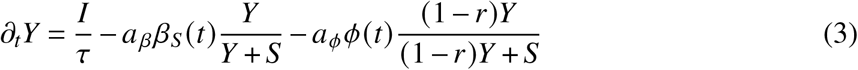

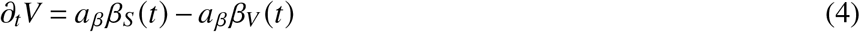

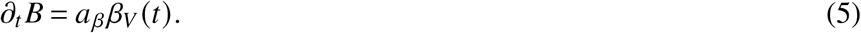

The last two equations are shown here for completeness, but note that the number of vaccinated and boostered individuals can simply be obtained from data, without integrating the dynamic equations, as their integrals can be evaluated analytically and are equal to the cumulative number of respective vaccinations. Above, *a*_*ϕ*_ and *a*_*β*_ are under-ascertainment ratios that account for infections and vaccinations that have not been reported. The time scale *τ* is equal to the average time after which an infected/recovered individual becomes eligible for reinfection or vaccination and 1 − *r* is the relative probability that an unvaccinated recovered person is reinfected as compared to a fully susceptible individual.

For our analysis, we draw 1,000 pairs of *a*_*ϕ*_ and *a*_*β*_ from shifted Gamma distributions with means ⟨*a*_*ϕ*_ ⟩ = 2, ⟨*a*_*β*_ ⟩ = 1.03, and standard deviations Std[*a*_*ϕ*_] = 1, Std[*a*_*β*_] = 0.02 that are bounded below by min(*a*_•_) = 1. Note that this distribution yields a median under-ascertainment ratio of *Q*_2_ [*a*_*ϕ*_] = 1.7, which is in line with results informed by seroprevalence data for Germany in 2020 [18]. Furthermore, with a 97.5th percentile of 4.7, the distribution is broad enough to account for occasional high under-ascertainment ratios that have been observed locally [10, 17, 18]. For infants, ascertainment is expected to be lower than for other age groups [21], which is why we double under-ascertainment ratios for this age group. We did not assume a higher under-ascertainment ratio for children older than 4 years, because regular screening via rapid antigen tests is mandatory in schools across the country [22]. We choose an eligibility time of *τ* = 90d, which is approximately of the same order as the time for antibody concentrations to decay after an infection [23]. While it falls in the lower bound of officially recommended time for recovered individuals to wait before getting vaccinated, surveys indicate that people might not strictly follow the official recommendation but get vaccinated earlier. Further, people with asymptomatic courses might have no knowledge about their infection, likely leading to a bias towards shorter times between infection and vaccination in those cases. The influence of lower and higher values of *τ* is investigated in a sensitivity analysis. The “recovered immunity” parameter *r* quantifies the relative efficacy against reinfection. For the Alpha variant, this efficacy was observed to be lower than the vaccine efficacy against infection by mRNA- or vector-vaccines [24], but of similar order as the vaccine efficacy against infection with Delta, taking on values of *r* ≈ 0.65 for both. As Omicron is considered to be a variant with partial immune escape, we set a lower default value of *r* = 1/2 for all variants, testing *r* = 0 (no protection against reinfection) and *r* = 1 (full immunity) in sensitivity analyses.

The daily vaccination rates *β*_•_(*t*) are obtained from data [2] and averaged over calendar weeks to remove weekly modulations. Likewise, infection rates of unvaccinated individuals *ϕ*(*t*) are obtained from reported data in the German reporting system SurvStat [25], which is available in aggregated form upon request. While the vaccination status is unknown for a substantial number of infections, we assume that for every day, the proportion of cases with unknown vaccination status that are, in fact, unvaccinated, is equal to the proportion of unvaccinated cases over the last seven days for which the vaccination status is known. This imputation method is performed for age- and region-stratified data.

For analyses disregarding infections with Omicron, we obtained the nation-wide and age-independent share of randomly sequenced samples in Germany [26] that the software framework “scorpio” identified as “Omicron” or “Probable Omicron” on a per-calendar-week basis by date of extraction (“Entnahmedatum”) as *σ*(*t*), assuming *σ*(*t*) = 0 for dates previous to Aug 1, 2021 and *σ*(*t*) = 1 for dates that exceed the last available date in the data. Then, all incidence rates were scaled as *ϕ*_*S*,pre−Omicron_(*t*) = *ϕ*_*S*_ (*t*) [1 − *σ*(*t*)]. Note that vaccination rates are unaffected by this procedure.

Population sizes stratified by age and state were requested from destatis [27].

Eqs. (1)-(5) are integrated using Euler’s method with Δ*t* = 1d until the last day of available incidence/vaccination data. For dates where data is unavailable, we assume the respective rates are equal to zero.

## III. RESULTS

We find an estimated nationwide median share of fully susceptible individuals of 7.0% (quartile range [3.6%–9.8%]). This result is, however, biased towards higher values due to a larger share of yet unaffected infants (44.6% [27.5%–56.8%]), children (22.5% [7.9%–34.3%]), and adolescents (5.0% [1.0%–10.1%]). For age groups that are associated with a higher probability of severe disease [28], we find a lower relative frequency of 3.5% [1.3%–5.5%] (adults), and 4.3% [2.7%– 5.8%] (elderly).

These values are achieved largely due to the (at the time of analysis still ongoing) Omicron wave. Ignoring infections with the Omicron variant, the nationwide age-independent share of fully susceptibles increases to 16.1% [14.0%–17.8%], i.e. Omicron infections are expected to have caused a reduction in fully susceptible individuals on the order of 10 percentage points at the time of writing, though this number differs by age group. While the change in relative frequency of fully susceptibles in the “adult” and “elderly” age groups was only about a few percentage points (median decreases from 9.2% to 3.5% and from 6.6% to 4.3%, respectively), the three youngest age groups were affected much more strongly, with median values of fully susceptible individuals dropping from 83.3% to 44.6%, from 63.5% to 22.5%, and from 23.8% to 5.0% with increasing age (cf. Fig. 2). If all variants are considered, the median share of fully susceptible “adults” and “elderly” barely differ (absolute difference of 0.8% points), likely due to a larger fraction of Omicron-recovered “adults” (Fig. 2).

**FIG. 1.**
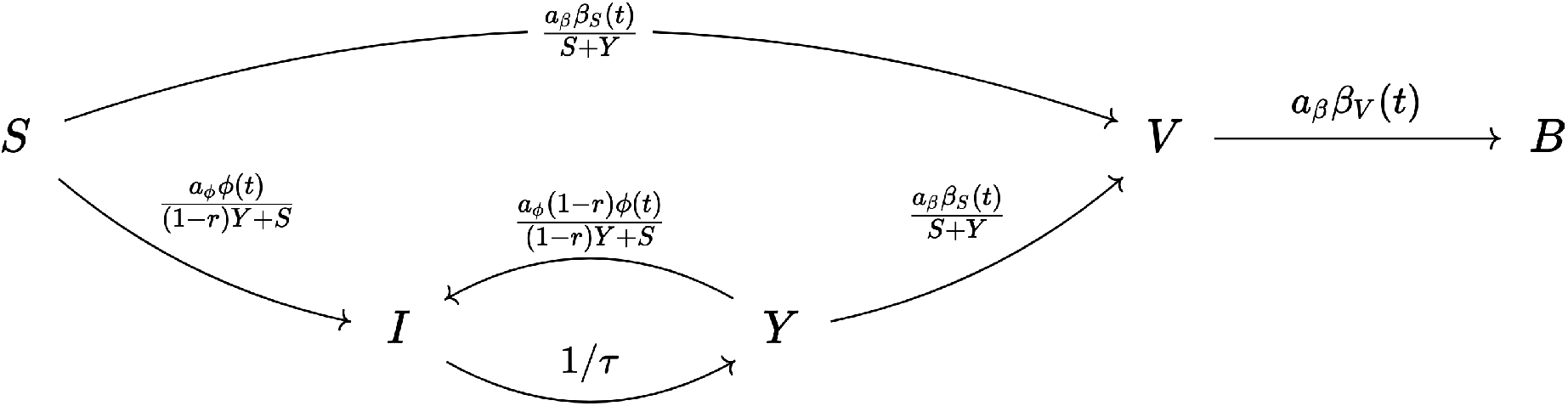
Simplified model schema. On each day, *a*_*β*_*β*_*S*_ (*t*)Δ*t* unvaccinated people become vaccinated, with under-ascertainment ratio *a*_*β*_ and Δ*t* = 1d. The probability that a newly vaccinated person has been infected before is proportional to the respective size of the subpopulation of recovered people that are eligible for vaccination *Y*. Furthermore, on each day, *a* _*ϕ*_ *ϕ*(*t*)Δ*t* unvaccinated people become infected, with underascertainment ratio *a* _*ϕ*_. The probability that a newly infected person has been infected before is proportional to the respective size of the subpopulation of recovered people that are eligible for reinfection (1 − *r*)*Y*, where 1 − *r* is the relative reinfection probability or “recovered immunity”. Recovered individuals are expected to reach eligibility for reinfection/vaccination after an average duration of *τ*. (Note that in the full model breakthrough and reinfections of vaccinated individuals are possible (see App. A 1).)

**FIG. 2.**
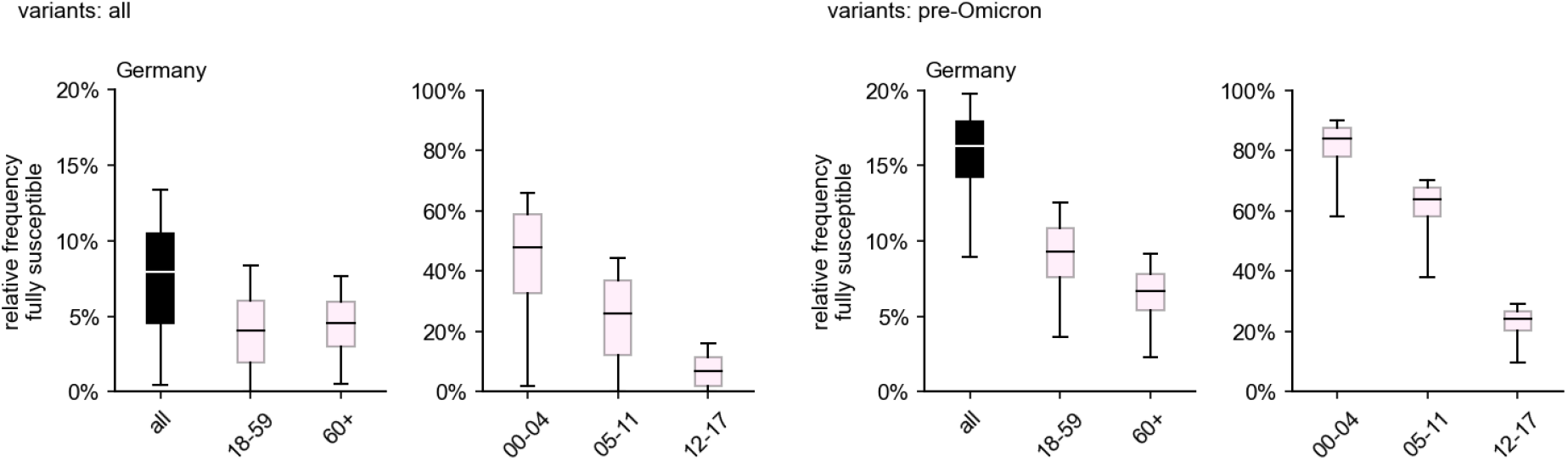
Estimated nationwide relative frequency of fully susceptible individuals by age group, considering vaccinations and infections that took place up to and including March 2022. Boxes represent the area between quartiles *Q*_1_, *Q*_3_ and whiskers the 2.5th and 97.5th percentiles, respectively, the median is shown as a horizontal line. (Left) Considering infections with any variant. (Right) Considering infections with any variant other than Omicron and its sublineages.

Although the relative frequency of fully susceptibles varies between federal state, certain commonalities are still shared. In all states, the frequency of fully susceptible individuals decreases with age, with a strong dependence on age for children. For ages 12-17, the frequency reaches values on the same order as those of the age groups “adults” and “elderly” (Fig. 3). Apart from the fact that adult and elderly age groups achieve relative frequencies of fully susceptible individuals below 10%, there are no other common patterns that stand out across all states regarding these age groups. In general, these age groups show overlapping quartile intervals, with the exception of Hamburg and Bremen, where “adults” show a comparatively lower relative frequency (Fig. 3). In fact, in Bremen virtually noone aged 18 and above is expected to not have been in contact with either virus or vaccine, according to the estimations.

**FIG. 3.**
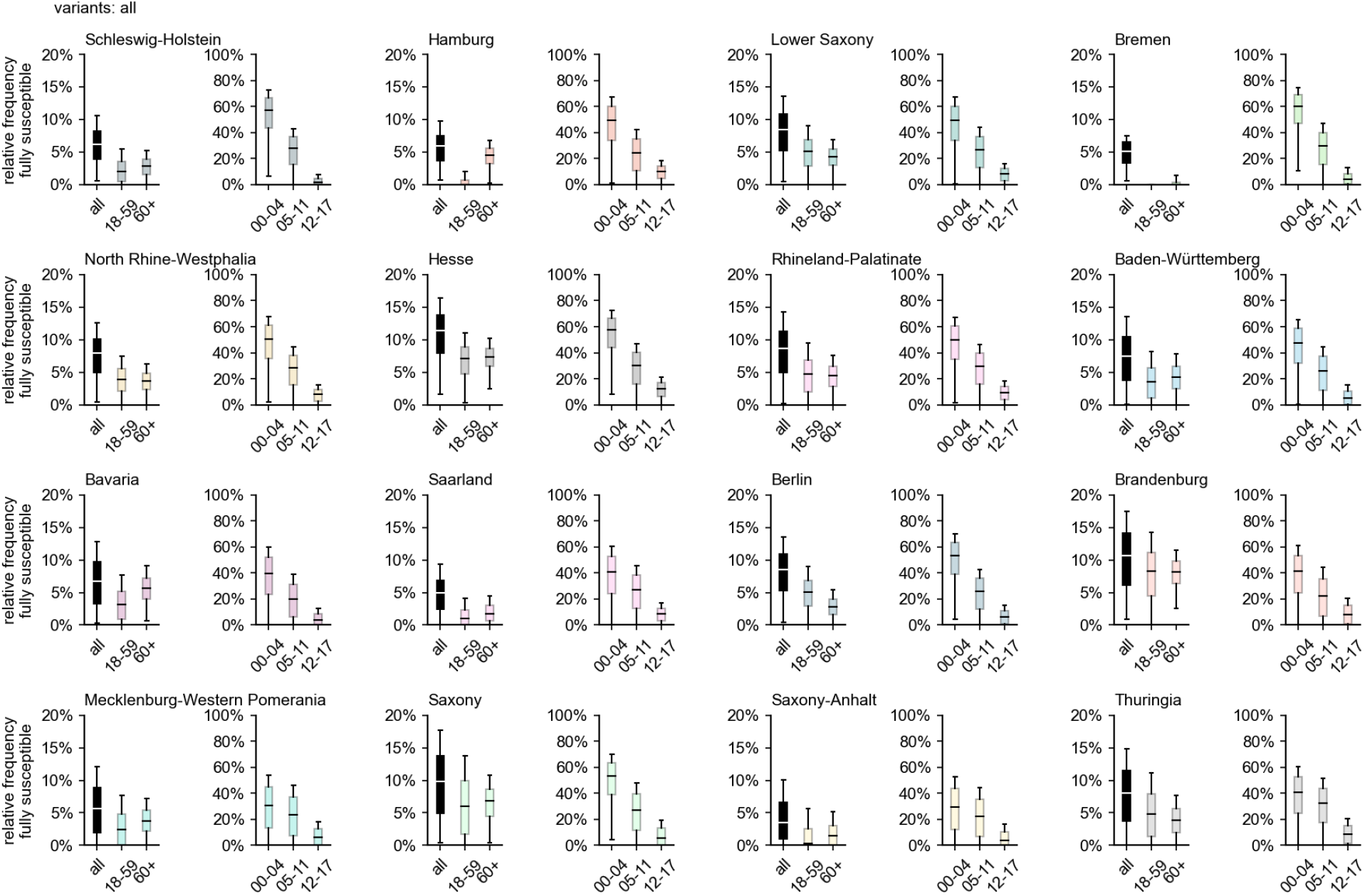
Estimated relative frequency of fully susceptible individuals by age group and region considering infections with any variant and vaccinations up to and including the Omicron wave (as of March 31, 2022).

In general, the above observations hold for the pre-Omicron analysis as well, except for the fact that, in the majority of states, the number of adults that were still unaffected decreased dramatically during the Omicron wave due to the large number of infections caused by the variant (comparing Figs. 4, 3). When excluding Omicron infections, the relative frequency of fully susceptibles differs across states on the order of ∼ 10%, with Saxony and Bremen as the states with largest (20.3%) and smallest (10.0%) respective median values of fully susceptible individuals (Fig. 4). Including infections with Omicron, the median range between states is reduced to a difference of 6.0% points (median of 10.7% in Hesse and 4.7% in Bremen).

**FIG. 4.**
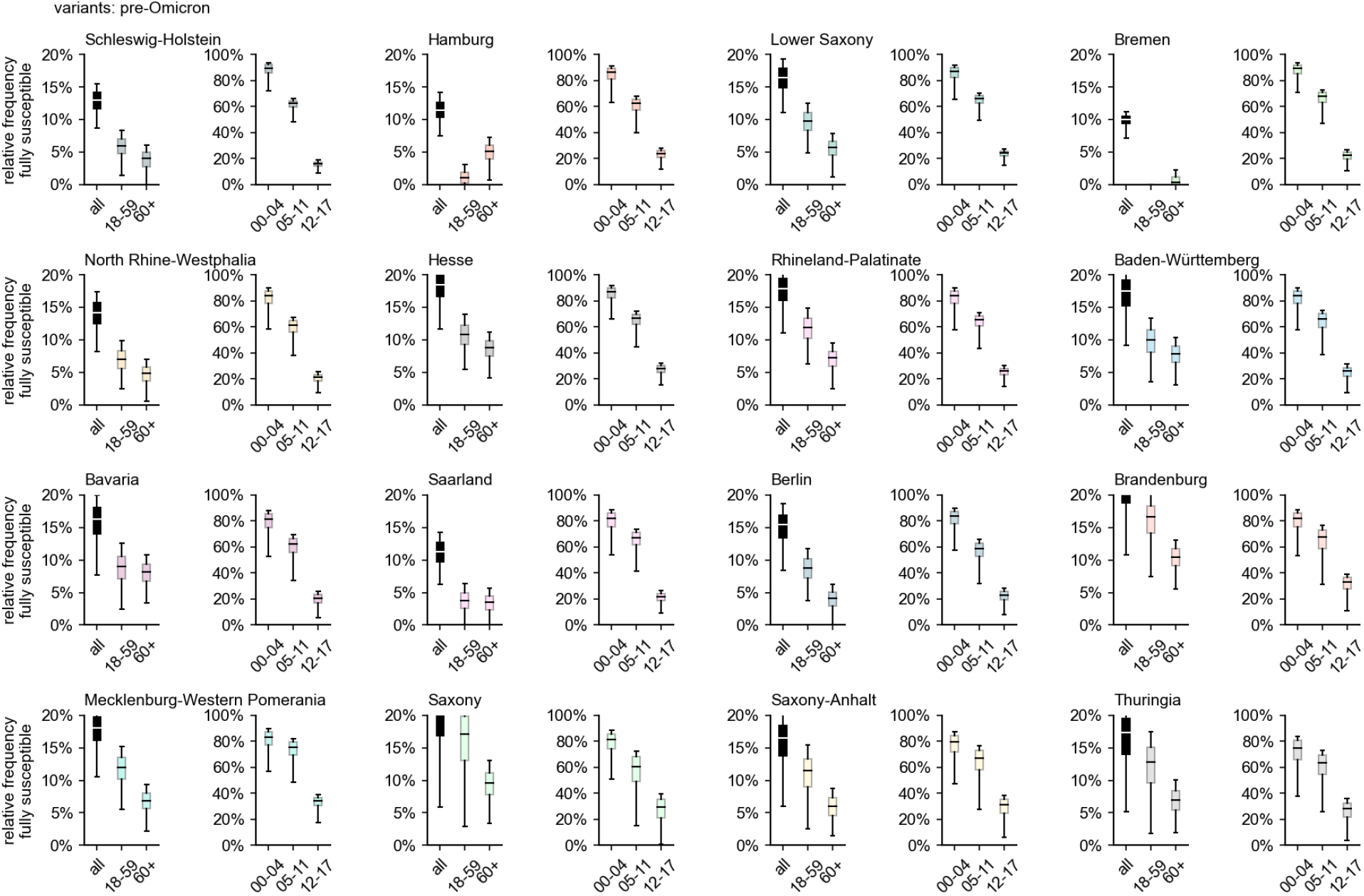
Estimated relative frequency of fully susceptible individuals by age group and region, disregarding infections with Omicron and its sublineages, based on data available up to and including March 2022.

Our results are robust against changes in assumed eligibility time *τ* and recovered immunity *r*, varying by a few percentage points in the nationwide average for all ages. For the most at-risk age groups, i.e. adults and the elderly, these results vary even less, indicating that the influence of these parameters decreases with age (see Sec. B and Fig. 7).

**FIG. 5.**
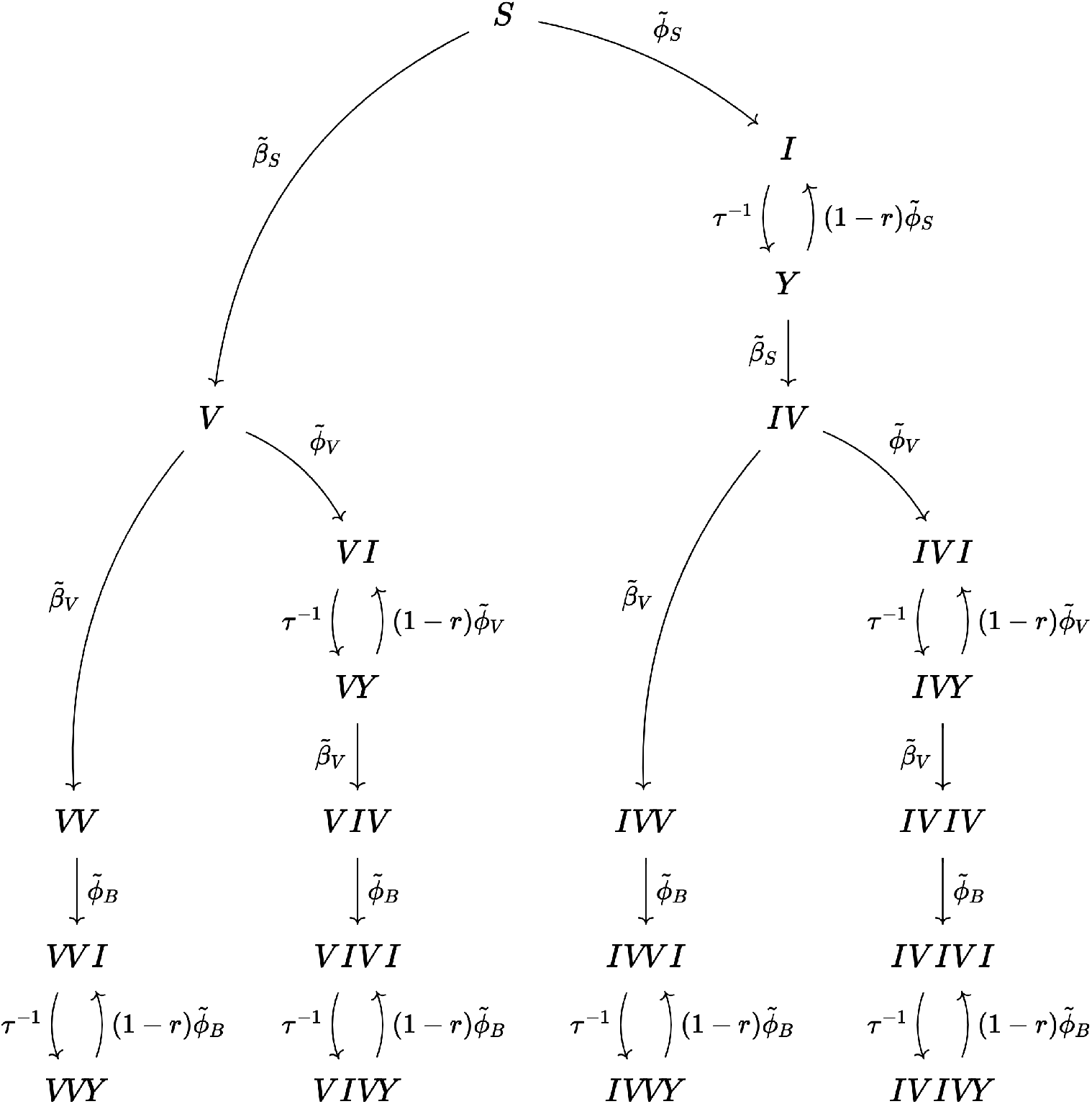
Vaccination/infection model given by Eqs. (A6)-(A36). Individuals can become infected and recover (compartments ending in *I*), vaccinated (compartments ending in *V*), or eligible for reinfection/vaccination after a previous infection after an average duration of *τ*^−1^ (compartments ending in *Y*). Initially, all individuals are susceptible (*S*). Transition rates are determined by data and scaled by assumed under-ascertainment ratios (not shown here). Individuals that are eligible for reinfection are associated with a relative reduction in susceptibility *r*. The order of *I* and *V* in individual statuses represent the order in which infections and vaccinations happened to the respective individuals.

**FIG. 6.**
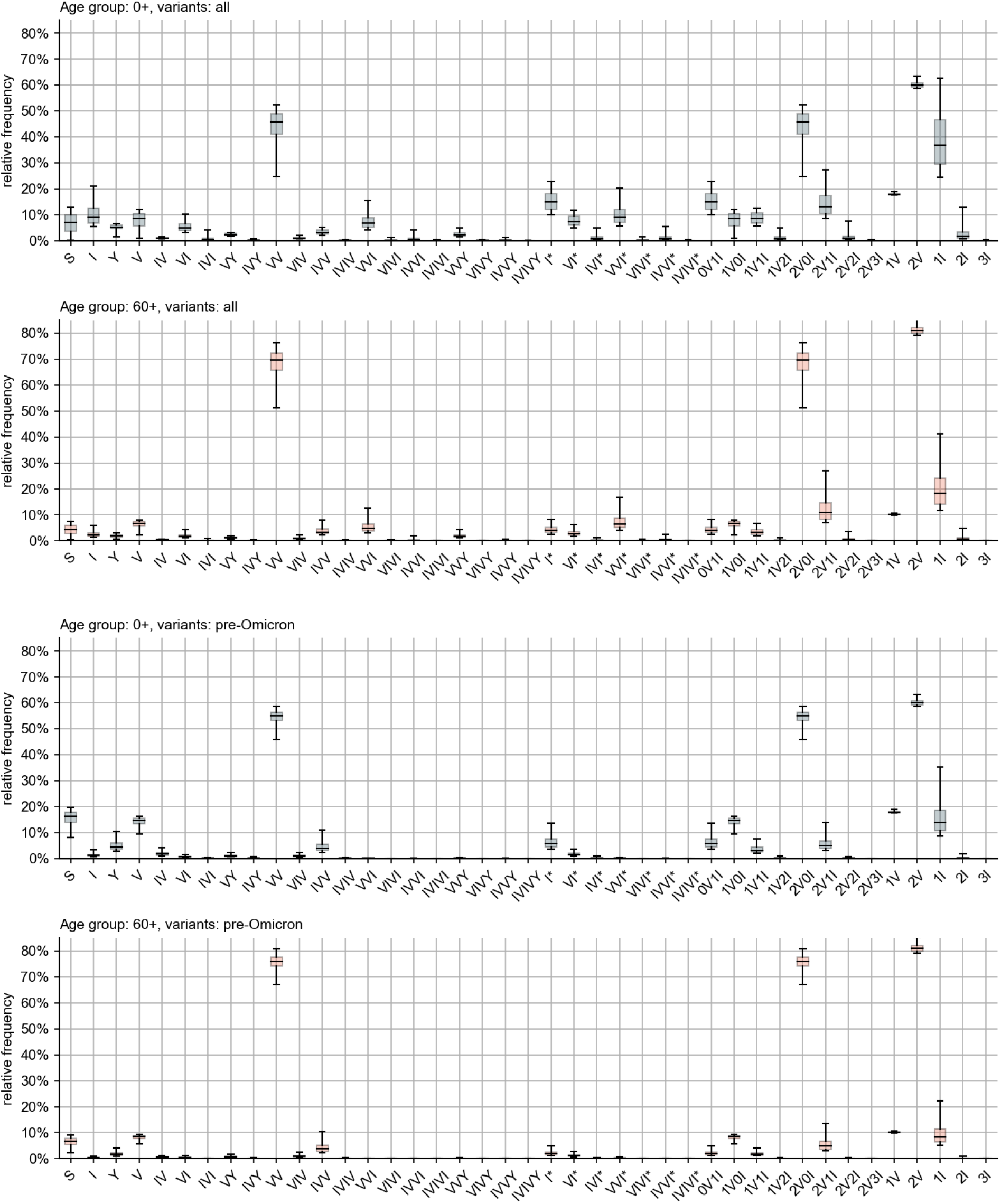
Relative frequency of all compartments given by vaccination and infection status across Germany, for all age groups and variants as well as for the elderly and pre-Omicron variants. Some compartments shown are aggregates, e.g. labels “*n*V*m*I” represent the number of individuals that were vaccinated *n* times and infected *m* times (re-infections excluded), labels “*n*V” give the number of individuals that were vaccinated *n* times, and labels “*m*I” are the number of individuals that were infected *m* times (re-infections excluded), see Eqs. ((A41))-((A61))

**FIG. 7.**
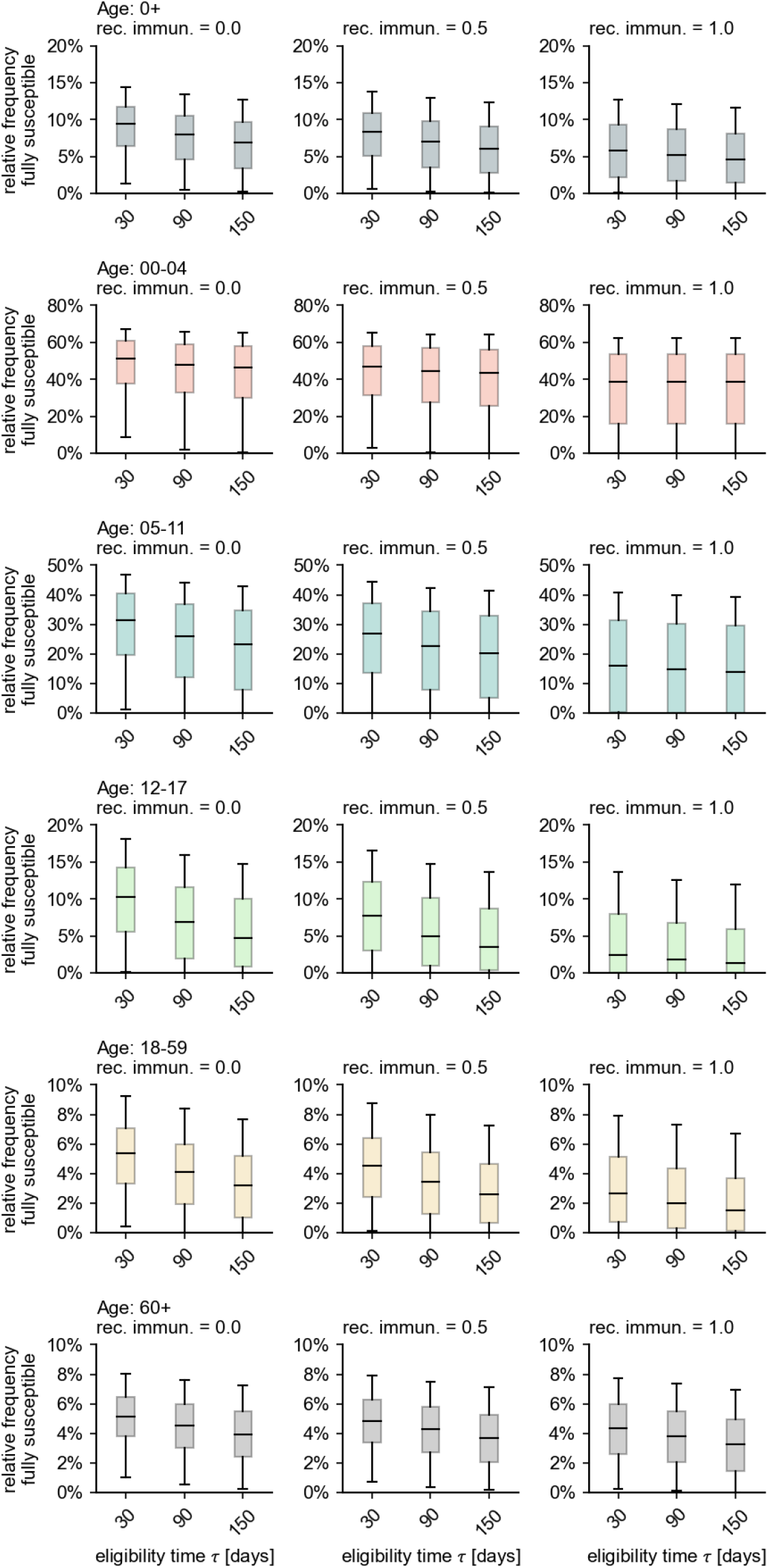
The influence of the assumed average eligibility duration as well as the long-term immunity of recovered individuals.

Regarding the detailed distribution of individuals by vaccination/infection status, we find that the largest single compartment of the model population is the group of people that has received a booster vaccination and has never been in contact with the virus (see Sec. B and Fig. 6), with unvaccinated recovereds comprising the second largest group. When first excluding, then including Omicron infections, both the number of non-infected vaccinateds and non-infected booster vaccinateds decreases by about 10 percentage points, demonstrating the relative efficacy of the booster vaccination against infections with the Omicron variant. The prevalence of compartments that count infected individuals decreases with the number of (breakthrough) infections per individual, which is unsurprising given that the model probability to become infected decreases exponentially with every new infection. Note that our model cannot, however, track the number of reinfections per individual between achieving the different vaccination statuses.

As under-ascertainment is expected to be larger for infants than for other age groups, we scaled the respective under-ascertainment ratio to always assume twice the value of other age groups. Because most children below 5 years of age will remain unvaccinated as per official recommendations, only infections reduce the number of fully susceptible individuals, and, therefore, the under-ascertainment ratio has a large influence (see Sec. B and Fig. 8). With the degree of underascertainment in this age group comparatively unclear, the results must be considered relatively uncertain for this age group.

**FIG. 8.**
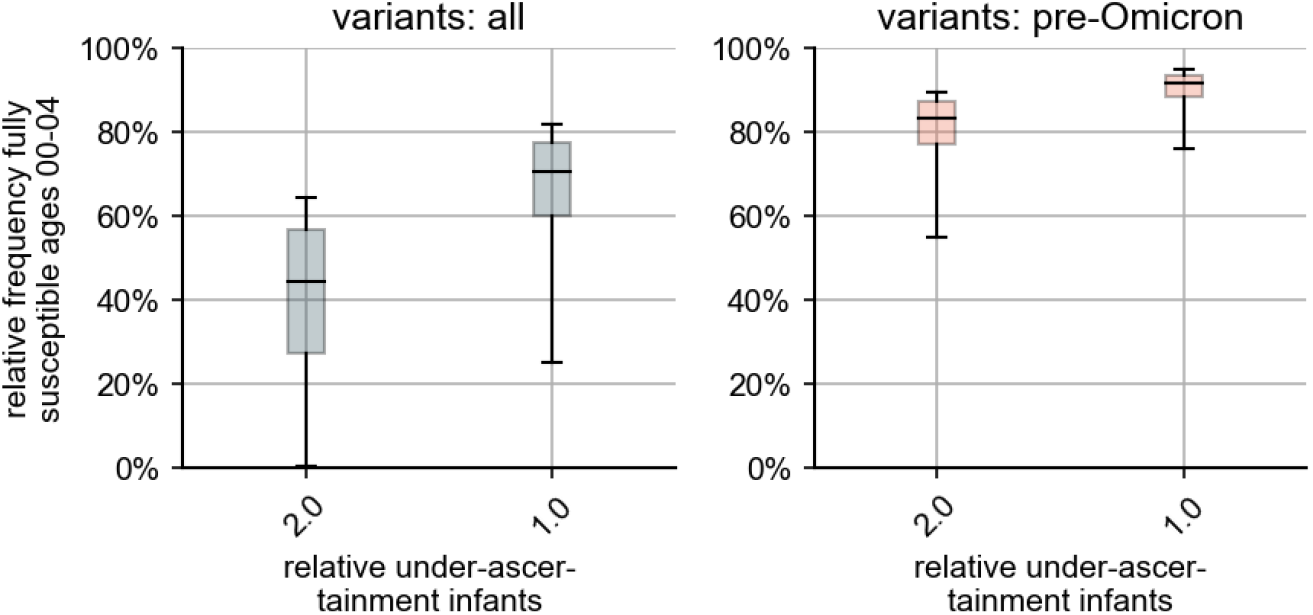
Influence of relative under-ascertainment for infants. For the main results, we assumed that the relative under-ascertainment factor assumes, for infants, a value of *a* _*ϕ*,infants_/*a* _*ϕ*_ = *ω* = 2. For *ω* = 1, the number of yet fully susceptible infants is higher than what we reported in the main text.

## IV. CONCLUSION & DISCUSSION

As the pandemic progresses, a central quantity that will determine the upcoming dynamics is the population-wide susceptibility against infection with known or future variants of SARS-CoV-2. While protection from infection, either derived from vaccination or natural infection, wanes over time and depends on the circulating virus variant, an estimation of the respective subpopulation sizes of people that suffered from (one or more) infections or were vaccinated/boostered gives valuable information about the size of the population that is, as of yet, still fully susceptible to infection, because these individuals are more prone to infection and severe disease as compared to vaccinated or recovered individuals, given that future variants do not fully escape this immunity.

Here, we found that in Germany, a nationwide single-digit percentage of individuals have not been in contact with either a variant of SARS-CoV-2 nor a vaccine against them, yet these results vary between regions and age groups. Despite the high number of reported infections in infants, children, and adolescents, a considerably high percentage of these age groups may still be fully susceptible to infection. This may become problematic if a variant emerges that causes more severe disease in these age groups than previous variants. Yet, we cannot rule out the possibility that we underestimated the extent of under-ascertainment in these age groups, as the factors we used where informed by seroprevalence studies based on blood samples donated by adults (ages 18–74), while it has been reported that under-ascertainment ratios can assume values ranging from 2 up to 6 or 8 for children [29–31].

In comparison, the age groups of adults and elderly showed a relatively low share of fully susceptible individuals, considering infections with all variants, on the order of 5%. Only considering infections with pre-Omicron variants, however, around 7.4%–10.7% of the adult population and 5.3%–7.8% of the elderly population may still be at risk of infection with variants that have a higher probability of causing severe disease than Omicron, potentially causing large outbreaks that could put high pressure on the public health system once again (with these numbers representing quartile ranges).

Our results are subject to a number of limitations and biases. For instance, the reported uncertainties (quartile ranges) are heavily determined by the choice of distribution of *a*_*ϕ*_. The distribution we chose has a median value of *Q*_2_ [*a*_*ϕ*_] = 1.7, which is slightly lower than what was observed in 2020 [18]. Moreover, the lower distribution bound of min(*a*_*ϕ*_) = 1 might be rather low, as such a value would mean that every infection has been reported, which is unlikely. Hence, at least the upper percentiles we report for *S*_∞_ might be overestimations. Furthermore, we assume the same distribution of under-ascertainment ratios for all German states, which might not reflect potential heterogeneities in local ascertainment particularly well.

Regarding modeling choices for the eligibility time, a short average duration after infection to be eligible for vaccination leads to larger proportions of vaccine-eligible people and, hence, to a higher overlap between the vaccinated and recovered subpopulations, thus increasing the estimated number of fully susceptibles. While we chose a comparably low value of 90d for this parameter, lower values cannot be ruled out. However, (i) the value we chose lies below the official recommendation, and (ii) changes in this parameter are not expected to change our results drastically, as was shown in a sensitivity analysis.

Likewise, shorter durations of eligibility for reinfection and lower values of long-term immunity of recovered individuals increase the likelihood that a reported infection of an unvaccinated individual was, in fact, a reinfection event, thus leading to higher values of fully susceptible individuals over all. As above, our results are robust towards variations in these parameters.

Regarding results on a regional level, reported vaccinations and infections might be skewed regionally when a large number of people live in one state but traverse to others to seek medical help. These considerations might explain the extreme results observed for Hamburg and Bremen, which are city states enclosed by others.

The last German census took place in 2011 and population sizes per age group and region have been imputed for the year 2020 based on this data, thus potentially being subject to over- or under-counting. Uncertainties in population size may introduce systematic errors on the order of a few percentage points in relative frequencies. When such a relative frequency reaches low values, these absolute errors on the order of a few percentage points can lead to high relative errors in the results.

Considering incidence rates, we imputed the total number of unvaccinated cases per day from cases with undetermined vaccination status by assigning them the “unvaccinated” status with probability proportional to the share of unvaccinated cases in the set of cases with determined status. This procedure can introduce systematic errors when the ascertainment of vaccination status is biased towards any of the vaccination states, which may occur, for instance, when the probability of status ascertainment increases with severity of disease. In this case, people with breakthrough infections may be less likely to have their vaccination status reported in the reporting system, which would mean that we overestimated the number of unvaccinated cases per day, introducing a bias towards lower values of the share of fully susceptible individuals.

For analyses regarding infections with variants prior to Omicron, we relied on the nationwide share of Omicron sequences, multiplying all incidence rates (regardless of region, age, or vaccine status) with this function. Since vaccines assume different efficacies against infection with different variants and will likely vary across ages and regions, this assumption is expected to introduce strong bias on a fine-grained population level, which may be expected to decrease when values are aggregated over regions or ages.

Our results cannot be used to predict the future course of the pandemic directly. In fact, since SARS-CoV-2 lacks phenotypical stability and neither infection nor vaccination elicit full long-term protective immunity, especially with respect to the prevention of infection and transmission, there are doubts that classical herd immunity can be reached for COVID-19 [32]. In several studies, hybrid immunity resulting from infection-acquired immunity boosted with vaccination conferred the strongest, or longer-lasting protection, respectively [33, 34]. Similarly, Omicron break-through infections in previously vaccinated individuals have been shown to drive cross-variant neutralization and memory B cell formation [35], suggesting that a combination of both, natural infection and vaccination, will have more impact on the future COVID-19 epidemiology than one of the events alone.

To sum up, our study shows that, presumably, only a small part of the German population has not yet been in contact with either a variant of SARS-CoV-2 or a respective vaccine against the disease they cause, up to and including March 2022. We show important proportions of fully susceptible elderly, who on average, by their age and age-associated morbidities, have a disproportionately elevated risk of severe disease. These shares differ by region and could motivate regionally targeted protection measures at the time of writing or in case of future outbreaks.

While the immunization campaign was successful in spring and summer 2021, in particular reaching a large proportion of vulnerable people, it thereafter had difficulties to completely close immunity gaps with vaccinations, albeit enhancing the protection of a large proportion of already vaccinated people with a large booster vaccination campaign by the end of 2021. Our results show that the Omicron wave had a high impact on naturally closing the aforementioned gaps. As mentioned above, however, having been in contact with a variant of SARS-CoV-2 is not a robust equivalent of immunity and may range from mild infection followed by rapid waning of antibodies and a highly uncertain degree of immunity, to a fully vaccinated status including a booster and a breakthrough infection, which confers a more long-lasting and robust degree of protection against severe disease. At the lower end of this spectrum of presumed immunity, our analyses show that one in six persons was never vaccinated but infected once or more, in the majority of cases with Omicron. This group faces higher uncertainties for the upcoming fall and winter since protection against severe disease may be more short-lived and too narrowly targeted to this variant.

## Data Availability

All data produced are available online at https://zenodo.org/record/6470799

## ACKNOWLEDGMENTS

The authors would like to thank Maria Waize and Matthias an der Heiden for helpful discussions. B.F.M. is supported as an *Add-on Fellow for Interdisciplinary Life Science* by the Joachim Herz Stiftung.

## Appendix A: Main model

### 1. Model formulation

We partition the population into *n*_*G*_ = 16 regions corresponding to the German states and *n*_*A*_ = 5 age groups corresponding to ages “00-04” (infants), “05-11” (children), “12-17” (adolescents), “18-59” (adults), “60+” (elderly), chosen in accordance with the population structure of publicly available vaccination data [2]. Consequently, for any region- and age-specific compartment *X*_*A,G*_, the nation-wide value is given as

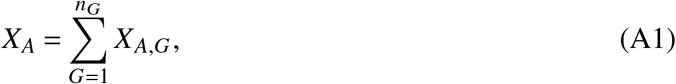

the corresponding value for all ages is given as

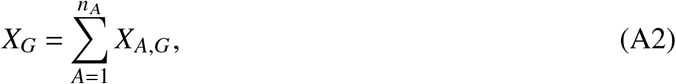

and the total value is

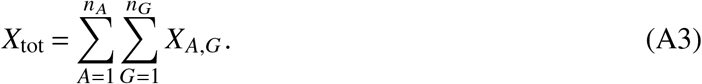

Because in the further analysis, none of the subpopulations are interacting, we will omit the region- and age-determining subscripts for simplicity.

For any population of size *N*, we are first and foremost interested in the number of susceptible individuals *S*, i.e. the number of individuals that have never been in contact with neither a variant of SARS-CoV-2, nor a vaccine against it. We assume that previous to the pandemic, no individual has had contact with any variant of SARS-CoV-2 or a vaccine against them, i.e. *S*(*t* = 0) = *N*. These susceptibles can then either (i) become infected (changing their status to *I*) or (ii) vaccinated (changing their status to *V*). The number of individuals changing their status per day is estimated from official data [2, 36], defining the number of reported newly infected unvaccinated individuals per day as *ϕ*_*S*_ and the number of newly vaccinated individuals per day as *β*_*S*_ (*t*). We obtain these rates on a calendar-week basis in order to remove weekly modulations. Because the vaccination status of new infections is unknown for a considerable amount of people, we impute *ϕ*_*S*_ from incomplete incidence data in a procedure outlined further below. The rates are to be interpreted in a way such that

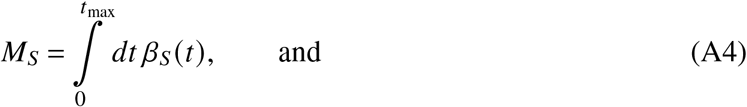

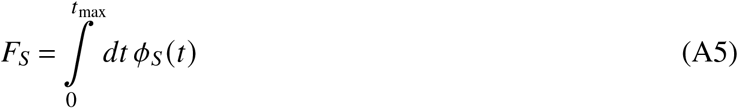

give the cumulative number of vaccinated individuals and the cumulative number of reported infections of unvaccinated individuals, respectively, both up to time *t*_max_.

At any time *t*, the number of individuals eligible to receive a vaccine is proportional to (a) the number of susceptible individuals and (b) the number of recovered individuals. We assume that infected individuals become eligible for vaccination after an average amount of time *τ* passes. Hence, after obtaining an infection, we assume that individuals change their status with rate 1/*τ* to become eligible (status *Y*). Then, the probability for a person that becomes vaccinated at time *t* to be of status *S* is given as *p*_*V,S*_ = *S*/(*S* +*Y*) and for status *Y* as *p*_*V,I*_ = *Y* /(*S* +*Y*). Consequently, the vaccination transition rate for both susceptibles and eligible recovereds to receive vaccination status is given as

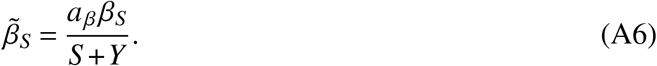

Here, we further introduced the under-ascertainment ratio of vaccinations *a*_*β*_. The corresponding transition processes are

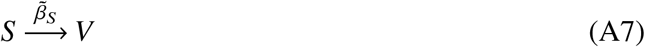

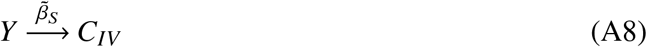

where *C*_*IV*_ represents the compartment counting individuals who became infected at least once before receiving a vaccination.

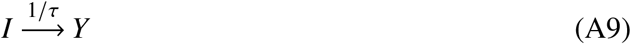

represents the process of recovered individuals becoming eligible for vaccination.

Similarly, the number of individuals eligible to transition to status “unvaccinated infected” is proportional to (a) the number of susceptible individuals and (b) the number of recovered individuals that are eligible for reinfection. We assume that individuals that recently suffered from an infection are fully immune, but may return to (partial) susceptibility after an average duration of *τ*, equating this to the average duration it takes to become eligible for vaccination for model parsimony and reasons outlined further below. Because reinfections are not registered in the German reporting system, we have to consider the relative probability for a recovered person to be reinfected by introducing an “immunity parameter” *r* that represents the relative probability of a recovered person to become infected after time *τ* since the last infection as compared to a fully susceptible person. Hence, the total number of people eligible to be counted as an infection of an unvaccinated individual at time *t* is given as *S* + (1 − *r*)*Y*, the probability that an unvaccinated person that becomes infected at time *t* has been infected before is *p*_*I,I*_ = (1 − *r*)*Y* /(*S* + (1 − *r*)*Y*), and *p*_*I,S*_ = *S*/(*S* + (1 − *r*)*Y*) that they have been fully susceptible. Consequently, the eligibility-corrected vaccination rate is given as

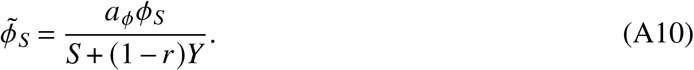

Here, *a*_*ϕ*_ is the under-ascertainment ratio, accounting for infections that have not been reported. The corresponding transition processes are

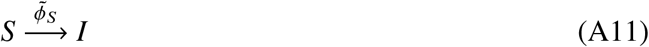

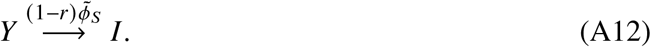

Again, Eq. (A9) represents the process of becoming eligible (both for vaccination after infection and reinfection).

Continuing with this line of argumentation, we further consider the adjusted rate of individuals that obtain a breakthrough infection as

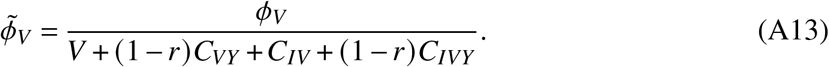

Here, *C*_*VY*_ are vaccinated individuals that suffered from a breakthrough infection before, and *C*_*IVY*_ counts individuals that, after recovery became vaccinated, then suffered from a breakthrough infection again. The respective transition processes are displayed in Fig. 5.

Similarly, the adjusted booster rate

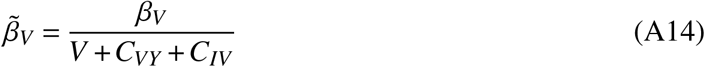

quantifies the rate with which previously vaccinated individuals receive a booster vaccination (processes shown in Fig. 5).

Finally, the adjusted booster breakthrough rate is

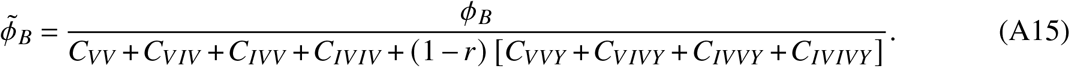

For every compartment *C*_•_, the order of *I* and *V* in the subscript • represents the order in which infections and vaccinations happened to the individuals counted in the respective compartment.

In total, the model is determined by the following set of ordinary differential equations (ODEs)

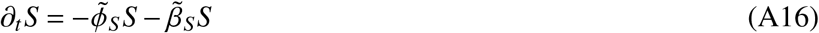

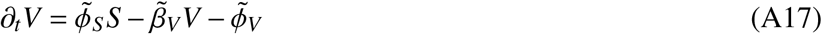

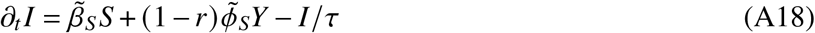

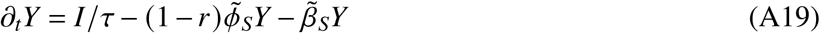

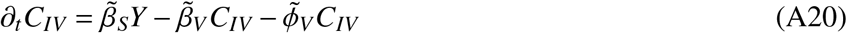

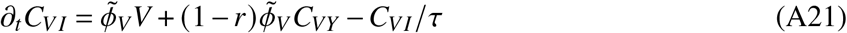

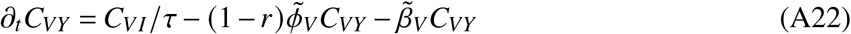

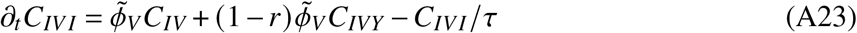

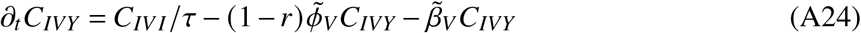

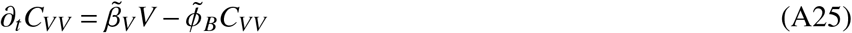

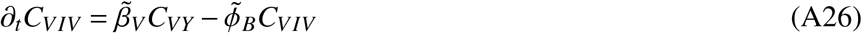

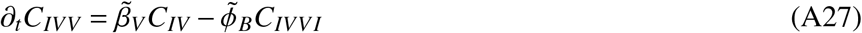

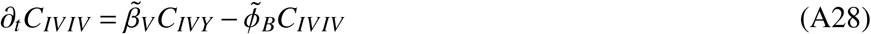

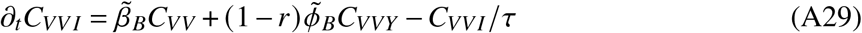

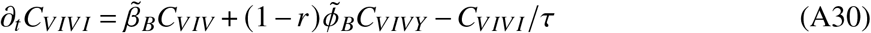

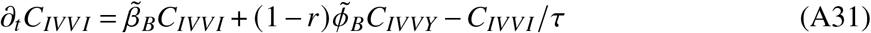

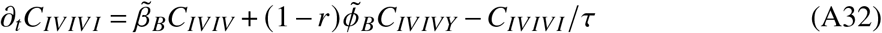

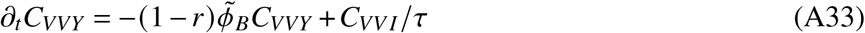

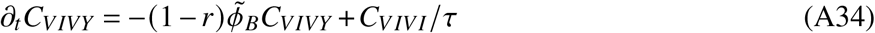

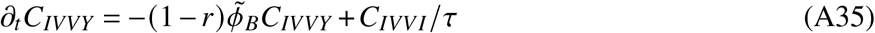

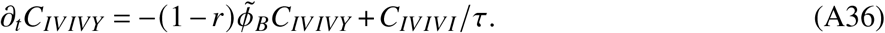

### 2. Parameters and data

#### a. Incidence by vaccination status

For each combination of age group and region, we obtain the daily number of reported new cases in unvaccinated 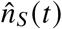 by “Meldedatum” (date of report), as well as the daily number of reported breakthrough infections 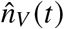, reported booster breakthrough infections 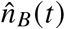, as well as the daily number of infections where the vaccination status is unknown 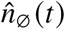 from the German reporting system SurvStat [25]. In order to assign vaccination statuses to cases where the status is originally unknown, we measure the proportion of infections per status in cases with known status in the last seven days and subsequently obtain the imputed number of daily cases as

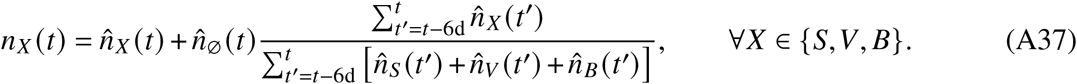

This procedure removes weekly modulations for the imputation. It might be biased towards any of the statuses *S,V, B* due to different probabilities of severe disease by vaccination status and thus of being reported in a system of primarily symptom-based testing. Note that, for no region and age groups there were days for which 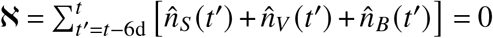 and 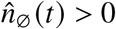, which is why we set 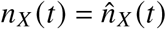 on days where ℵ = 0. With the above definition, the infection rates are given as

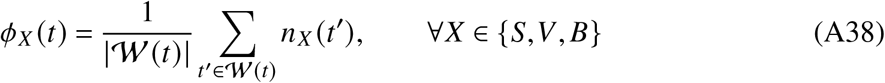

where 𝒲 (*t*) is the set of days *t*^′^ in calendar week of day *t* meeting *t*^′^ < *t*_max_.

#### b. Vaccination rates

Similarly, weekly vaccination rates are given as

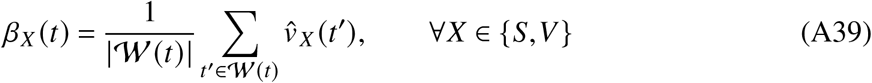

with 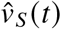 and 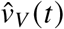 being the number of new vaccinations (new booster vaccinations, respectively) on day *t*. We define “new vaccinations” as entries in the data provided in [2] that have an “Impfschutz”-field value of “2”, and as “new booster vaccinations” as entries that have an “Impfschutz”-field value of “3”, ignoring single-shot vaccinations with value “1” (in the data, confirmed recovered individuals that received a single vector- or mRNA-vaccine dose are counted as being fully vaccinated with an “Impfschutz”-field value of “2”). The share of the population that received only one dose of an mRNA or the Vaxzevria vaccine is expected to be on the order of 1% of the German population up to and including March 2022 [2]. In the model, the infection of these individuals follows the same dynamics as the infection of fully susceptible individuals. Hence, ignoring this vaccination state will barely affect the results.

Note that we ignore the small number of vaccinations associated with the region “Bund” (region id “17”).

#### c. Under-ascertainment

Based on seroprevalence data collected over the first waves in Germany, a nation-wide underascertainment ratio of *a*_*ϕ*_ ≈ 2 was found, with regional variations that went up to a factor of *a*_*ϕ*_ ≈ 5 in regions of large outbreaks [10, 18]. In absence of more fine-grained and temporally resolved estimations, we assume an under-ascertainment of *a*_*ϕ*_ = 1 + *â*_*ϕ*_ with *â*_*ϕ*_ being a Gamma-distributed random variable such that ⟨*a*_*ϕ*_ ⟩ = 2 and Std[*a*_*ϕ*_] = 1.

It has further been reported that there might be low under-ascertainment in vaccinations [37]. We assume an under-ascertainment of *a*_*β*_ = 1 + *â*_*β*_ with *â*_*β*_ being a Gamma-distributed random variable such that ⟨*a*_*β*_ ⟩ = 1.03 and Std[*a*_*β*_] = 0.02.

Infants are less likely to display symptoms when infected and are not subject to the strict testing strategies applied in schools [38]. A lower ascertainment in this age group is, therefore, a plausible assumption. We hence assume double the value of the under-ascertainment ratio for this age group.

#### d. Eligibility time and immunity of recovered individuals

We assume an average eligibility time of *τ* = 90d for vaccination after infection or reinfection. Regarding reinfection, this is a reasonable time scale, as it is of the order of the mean duration neutralising antibodies can be found after an infection. For vaccinations, the official assumption for receiving a vaccine after infection has been 3–6 months. In non-representative survey data, it was found that participants generally followed these recommendations, with a large number of participants waiting less and became vaccinated about 3 months after a confirmed infection. While the cohort of this study is assumed to be composed of highly compliant individuals, the average time to receive a vaccination is also lowered assuming a large number of asymptomatic infections, where the date of the infection might be unknown to recovered individuals themselves. Note, however, that we test the influence of this parameter on our results in a sensitivity analysis (see App. B).

We recognize that recovered individuals might still have a lowered susceptibility for reinfection even after transitioning to the eligibility state. The “recovered immunity” parameter *r* quantifies the relative efficacy against reinfection. For the Alpha variant, this efficacy was observed to be lower than the vaccine efficacy against infection by mRNA- or vector-vaccines [24], but of similar order as the vaccine efficacy against Infection with Delta, taking on values of *r* ≈ 0.65 for both. As Omicron is considered to be a variant with partial immune escape, we set a lower default value of *r* = 1/2 for all variants, testing *r* = 0 (no protection against reinfection) and *r* = 1 (full immunity) in sensitivity analyses.

#### e. Variant share

For analyses disregarding infections with Omicron, we obtained sequences that were sampled randomly nation-wide and independent of age [26]. For each calendar week *w* we obtained the total number *m*(*w*) of randomly sampled sequences with date of extraction *t* that lie in *w*. We further aggregated the number *m*_*o*_ (*w*) of randomly sampled sequences that the software framework “scorpio” identified as “Omicron” or “Probable Omicron”. Then, the share of Omicron on day *t* is given as

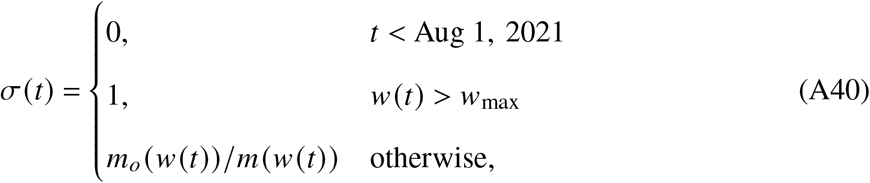

with *w*_max_ being the last week for which data was available.

For analyses labeled “pre-Omicron” we analyzed the model with all incidence rates being scaled as *ϕ*_•,pre−Omicron_(*t*) = *ϕ*_•_(*t*) [1 − *σ*(*t*)].

#### f. Simulations

We draw 1,000 pairs of (*a*_*ϕ*_, *a*_*β*_) as described above and assume those under-ascertainment ratios to be constant across all respective ages and regions (bar infants, whose under-ascertainment ratio is set as *a*_*ϕ*,infants_ = *ωa*_*ϕ*_ with *ω* = 2 to account for the fact that under-ascertainment is expected to be higher in this age group). Then, Eqs. (A16)–(A36) are integrated with Euler’s method using a time step of Δ*t* = 1d, starting on Jan 6, 2020 until March 31, 2022. We then obtain the final state of the compartments, and additionally aggregated states as

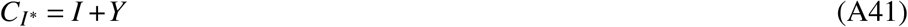

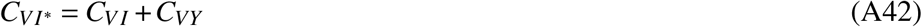

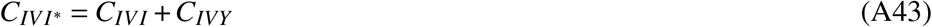

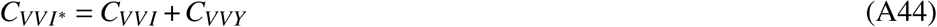

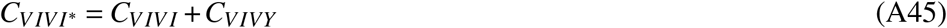

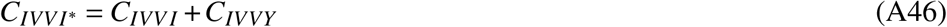

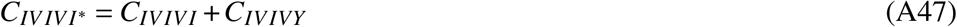

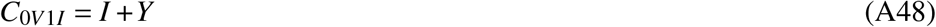

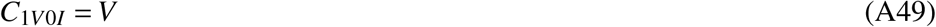

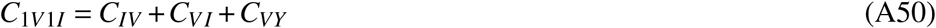

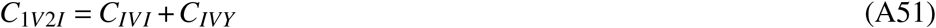

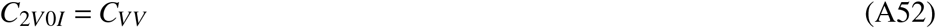

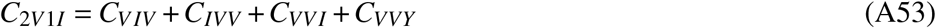

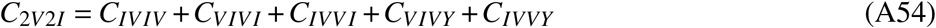

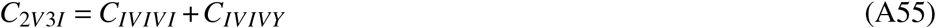

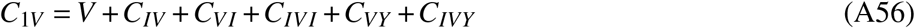

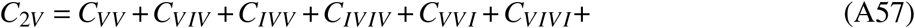

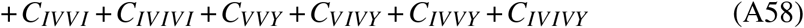

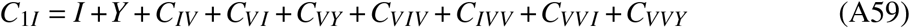

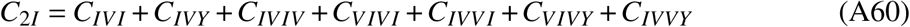

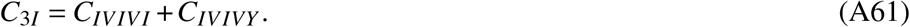

These states combine compartments that have certain commonalities, e.g. compartments *C*_*nVmI*_ is the number of individuals that were vaccinated *n* times and infected *m* times (re-infections excluded), *C*_*nV*_ is the number of individuals that were vaccinated *n* times, and *C*_*mI*_ is the number of individuals that were infected *m* times (re-infections excluded, which means that if an individual was infected *m* = 3 times, they must have been infected before, between, and after the respective inoculations.

We test how robust our results are if per region and age group, individual pairs (*a*_*ϕ*_, *a*_*β*_) were drawn from their respective distribution, i.e. assuming heterogeneous under-ascertainment in ages and regions per simulation run, which could potentially change the width of the distribution of respective aggregated values, finding that it does not have a substantial effect.

The results of these simulations can be obtained from [39].

## Appendix B: Sensitivity and other analyses

Nation-wide results for all compartments as well as Eqs. ((A41)–(A61)) can be found in Fig. 6. The compartment with the largest share of the population is *C*_*VV*_, i.e. boostered and never infected, assuming a value of 45.8% [41.1%–49.0%]. Considering all variants, the second largest value can be found for individuals that have never been vaccinated but infected once or more with *C*_*I*_* assuming 14.9% [12.0%–18.1%]. This value is considerably lower (5.6% [4.3%–7.5%]) when infections with Omicron are excluded. Likewise, the share of vaccinated, yet non-infected individuals *V* is estimated to assume 14.6% [13.4%–15.3%] with Omicron infections excluded, but 8.6% [5.8%–10.6%] considering all variants. With Omicron infections excluded, the boostered and non-infected population assumes an estimated size of 54.9% [53.1%–56.2%], demonstrating the increased efficacy of the booster vaccination against infection with Omicron as compared to individuals who only finished the first vaccination series.

Regarding the influence of eligibility time, higher values lead to a lower probability of reinfections and vaccinations of recovereds during the most active period of the vaccination campaign, implying the estimated number of fully susceptible individuals decreases with increasing *τ*. Likewise, the assumed immunity of recovereds *r* leads to a decreasing value of fully susceptible individuals. The results we reported above lie central within the range of results for extreme value pairs of *τ* = 30d, *r* = 0 (low), as well as *τ* = 150d, *r* = 1 (high). For instance for all ages, the results vary between median values of 9.5% (low) and 4.6% (high) with our reported result in the main text (*τ* = 90d, *r* = 0.5) being equal to 7.0%. The influence of these parameters are higher for the younger population with a “low”-to-”high” variation leading to respective median ranges of 51.0% to 38.8% (infants), 31.5% to 14.0% (children), and 10.3% to 1.3% (adolescents). In the older population, the influence of these parameters is rather small, leading to median ranges of 5.4% to 1.5% (adults) and 5.1% to 3.3% (elderly). These results are displayed in Fig. 7.

In the main text, we assumed that the relative under-ascertainment factor in infants assume a value of *a*_*ϕ*,infants_/*a*_*ϕ*_ = *ω* = 2. For *ω* = 1, fully susceptible infants is higher than what we reported in the main text (see Fig. 8. Since empirical values for *ω* are difficult to obtain, we are probably underestimating the uncertainty in our results for infants.

## Appendix C: Additional, sophisticated Model

We further want to develop a model that allows waning to be included in the analyses and could therefore potentially be used to estimate seroprevalence in future studies.

We hypothesize that exposure to either the pathogen or a vaccine results in an initial immune response that then decays over a period of time and account for this by introducing intermediate compartments representing different gradations of immunity.

We define as *S* susceptibles, *I* infected, *V* vaccinated, *Y* breakthroughs from vaccinated *V* and *U* as breakthroughs from boostered *B*. For each compartment *X*, we consider *n*_*X*_ + 1 gradations, i.e. we assume that individuals who reach the status *X* pass through intermediate compartments in the form of a chain from initial *X*_0_ to final *X*_*n,X*_, per transition *X*_*i*_ → *X*_*i*+1_ with transition rate 1/*τ*_*X,i*+1_. This means that for each individual, each of these transitions is subject to a random delay

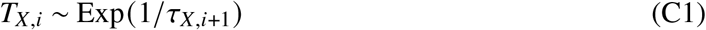

where Exp(*λ*_*X*_) is an exponential distribution with mean 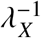. This approach allows us to more accurately model both waning of immunity and the timing of vaccination or breakthrough infection. For susceptibles, we set *n*_*S*_ = 0, i.e. no transitions and exactly one gradation.

We denote 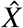 as the total number of individuals in status *X* that are susceptible to infection. That is, we define

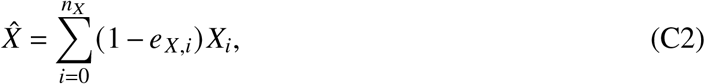

where *e*_*X,i*_ is the susceptibility reduction of a person in status *X*_*i*_ (due to previous infection or vaccination).

We define 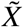 as the total number of individuals in status *X* who can receive one or the next vaccination. Usually, this is the case after a defined time Θ_*X*_ has passed since the last infection or the last receipt of a vaccine dose (comparable to the ‘eligibility time’ used in the main analyses of this study). The total time it takes for an individual in status *X*_*i*_ to reach status *X*_*i*+1_ is given by the random variable

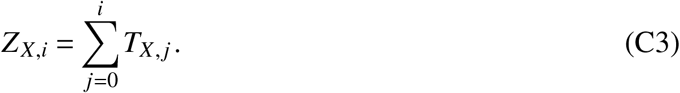

Let *F*_*X,i*_ (*z*) be the cumulative distribution function of the random variable *Z*_*X,i*_. Then, the probability *w*_*X,i*_ that a given individual in status *X*_*i*_ has been in status *X* for longer than Θ_*X*_ is given by

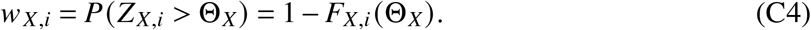

We find such

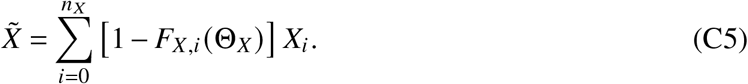

The probabilities *w*_*X,i*_ = 1 − *F*_*X,i*_ (Θ_*X*_) are constant and can thus be determined numerically after defining the times {*τ*_*X,i*_} and Θ_*X*_. For susceptibles, let 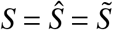.

Let ℐ(*X*) be the compartment to which an individual in status *X* transitions after infection and 𝒱(*X*) be the compartment to which an individual in status *X* transitions after vaccination. We define the following transitions

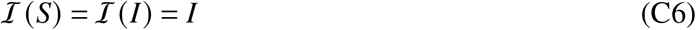

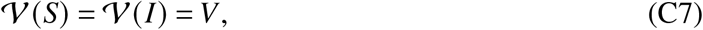

i.e. susceptibles *S* who become infected transition to status *I* and susceptibles who are vaccinated transition to status *V*. Recovered *I* who become infected again transition to status *I* and recovered people who get vaccinated transition to status *V*. Furthermore,

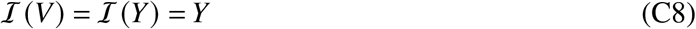

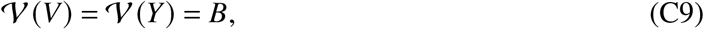

i.e. vaccinated individuals *V* who become infected transition to status *Y* and those vaccinated that receive a third dose transition to status *B*. Breakthrough-recovereds *Y* who become reinfected again transition to status *Y* and breakthrough-recovered individuals who become vaccinated transition to status *B*. Last,

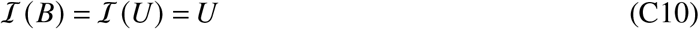

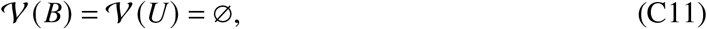

i.e. boostered persons *B* who become infected transition to status *U* but further vaccination is not provided. Recovered booster vaccinated persons *U* who become infected again will again transition to status *U*. The dynamics of all states *X*_*i*_ follows

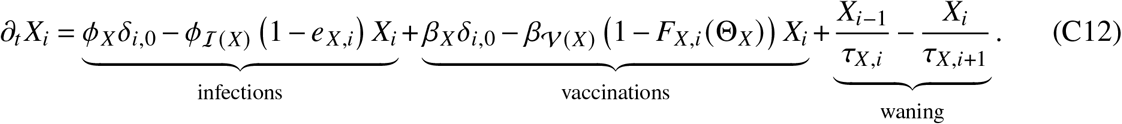

By definition, we have *X* _*j*_ = 0∀ *j* < 0 ∧ *j* > *n*_*X*_ + 1, as well as *ϕ*_∅_ = 0 and *β*_∅_ = 0. Furthermore, we set *β*_*S*_ = *β*_*I*_ = *β*_*Y*_ = *β*_*U*_ = 0 and *ϕ*_*S*_ = *ϕ*_*V*_ = *ϕ*_*B*_ = 0, that is, there are no infections ending in vaccination compartments and no vaccinations ending in infection compartments and no transitions ending in *S*. Additionally, susceptibles are maximally susceptible (i.e. *e*_*S*_ = 0) and from *n*_*S*_ = 0 follows *w*_*S*_ = 1. To ensure the validity of transition terms in intermediate compartments, we additionally define *τ*_*X, j*_ ≠ 0∀*X, j* ≤ 0 ∧ *j* > *n*_*X*_ + 1.

With regard to under-reporting, we assume that under-ascertainment ratios are already included in the respective rates *ϕ*_•_ and *β*_•_.

Finally, the aim of this analysis is to estimate seroprevalence at time *t*. For each state *X*_*i*_ ≠ *S*, we denote by *p*_*X,i*_ the probability that antibodies are found in a person in state *X*_*i*_. Then, the seroprevalence *P* of the age group/population of consideration is given as

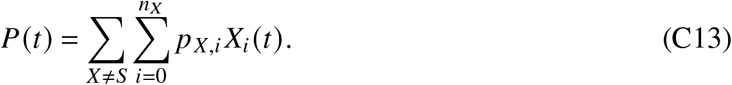

The model is illustrated in Fig. 9.

**FIG. 9.**
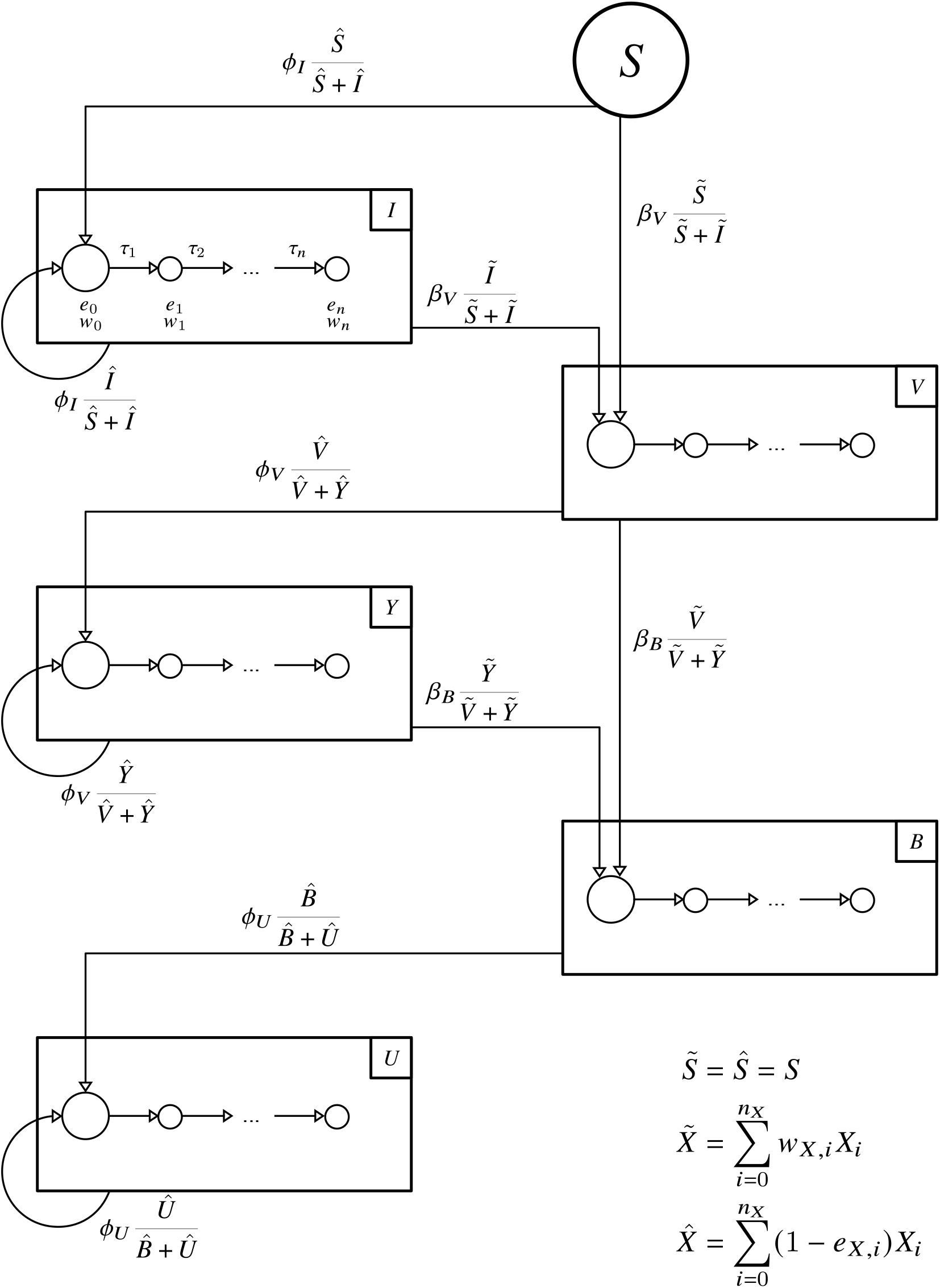
Detailed model that includes waning.

A large number of parameters are required to calibrate the model. For each state *X* ∈ {*I,V, Y, B,U*} the number of transitions *n*_*X*_ have to be defined, then *n*_*X*_ mean transition times as well as *n*_*X*_ + 1 susceptibility reductions. For compartments *I,V, Y* and *B*, eligibility times Θ_•_ for receiving a vaccination are to be determined. From reporting data, we obtain the daily number of new infections of unvaccinated *ϕ*_*I*_ (*t*), vaccinated *ϕ*_*V*_ (*t*) and boostered *ϕ*_*U*_ (*t*) individuals. From the vaccination archive, we obtain the daily number of completed initial vaccination series *β*_*V*_ (*t*) and booster vaccinations *β*_*B*_ (*t*). Under-reporting of infections and booster vaccinations must be estimated and accounted for in the respective rates. For each state *X*_*i*_ ≠ *S*, the probability *p*_*X,i*_ of finding antibodies in a person in state *X*_*i*_ must also be defined.

All these parameters have to be determined for each of the subpopulations (age groups, regions).

